# High tumor mutational burden predicts survival and responses to ICI immunotherapy in a cancer-dependent manner

**DOI:** 10.1101/2023.08.29.23294599

**Authors:** Feng Tang, Tian Lan, Zhen-Yuan Liu, Ze-Fen Wang, Zhi-Qiang Li

**Affiliations:** Brain Glioma Center & Department of Neurosurgery, Zhongnan Hospital of Wuhan University, Wuhan, Hubei, China; Department of Physiology, Wuhan University School of Basic Medical Sciences, Wuhan, Hubei, China

**Keywords:** Tumor mutational burden, immunotherapy, immune checkpoint inhibitors

## Abstract

**Objective:** High tumor mutational burden (TMB) is a promising biomarker for patients with immunotherapy in certain types of solid tumors. This article focuses on exploring possible universally optimal cutoffs of TMB for predicting immune checkpoint inhibitors (ICIs) response and prognosis for eight types of cancers.

**Methods:** The present study collected eight types of tumors including 2767 patients receiving immune checkpoint inhibitors (ICIs) treatment and 14862 patients without ICI treatment. We tried to explore optimal cutoffs of TMB in each type of tumor via selecting several possible cutoffs of TMB including 10mut/Mb, top 10%, 20%, and 30% of TMB within each histology.

**Results:** We found that there was a significant difference in TMB values between ICI-treated and non-ICI-treated groups. The cutoff of TMB appropriate for predicting response rates, progression rates, and survival rates was varied in ICI therapy patients. Moreover, the optimal cutoff of TMB for predicting progression-free survival and overall survival in different types of the tumor was also quietly different.

**Conclusion:** Our current study suggested that TMB predicts prognosis and responses to ICI treatment in a cancer-dependent manner.

## 1. Introduction

In recent years, the use of immune checkpoint inhibitors (ICIs), such as those targeting the programmed death 1(PD-1)/ ligand of the PD-1(PD-L1) axis and cytotoxic T lymphocyte antigen 4(CTLA4) have revolutionized cancer treatment for the improvements in overall survival across multiple solid malignancies [1–5]. Although nearly 45% of US cancer patients are eligible for ICIs therapy, the percentage of patients who benefit from this therapy was only 12.45%[6]. Therefore, the identification of predictive biomarkers of ICIs response is vital for the selective use of ICIs and the evaluation of possible mechanisms of immunotherapy resistance.

Accumulation of somatic mutations is the main cause that contributes to tumor development. Increased tumor-specific somatic mutations enable tumors to express more neo-antigen on the cell surface, which could be recognized by the immune system. Eventually, these immunogenic tumors exhibit high sensitivity to immunotherapy [7, 8]. Tumor mutational burden (TMB) is defined as the total number of somatic mutations per coding area of a tumor genome. It is a measure of the number of mutations in cancers. Patients with higher TMB generally harbor more neo-antigens to trigger a T-cell response, which therefore increases chances for tumor cell eradication[9]. Emerging evidence has shown that high TMB is a leading candidate biomarker and correlates with the clinical benefit of ICI therapy [10–13]. However, a systemic bioinformatics analysis of the landscape of mutation burden across cancer types containing 100,000 cancer cases demonstrated that TMB was very heterogeneous, with a range of 0–1241mut/Mb across cancer types[14]. Since the TMB distribution is in a cancer-type-dependent manner, an open question is how to determine optimal TMB cutoffs suitable for evaluating ICI therapy response in given tumor types. Morris et al. defined TMB subgroups by percentile within each histology and the highest 20% in each histology was used for cutpoints. However, the association between TMB (top 20%) and overall survival was only found in that research in four out of ten tumor types [15].

In 2020, the United States Food and Drug Administration (FDA) approved pembrolizumab (one kind of PD-1 inhibitor) for the immunotherapy of unresectable or metastatic adult and pediatric solid tumors of any histologic type with TMB ≥10 mut/Mb. This approval was based on the finding of a high objective response rate and overall survival among 102 patients with TMB who were treated with pembrolizumab monotherapy in the Keynote-158 trial containing ten tumor types[16]. The question remains whether TMB≥10mut/Mb also applies to universal ICI therapy instead of only pembrolizumab; and whether TMB≥10mut/Mb is an appropriate biomarker for other tumors (beyond tumor types included in the Keynote-158 trial) receiving ICI treatment.

In the present study, two ICI cohorts (total 2767 cases) and one non-ICI cohort (total 14862 cases) comprised of eight types of tumors were included. We investigated the correlation between TMB cutoffs and response rates, progression rates, progression-free survival (PFS) as well as overall survival (OS). We also tried to explore optimal cutoffs of TMB for each type of tumor via selecting several possible cutoffs of TMB including 10mut/Mb, top 10%, 20%, and 30% of TMB within each histology.

## 2. Methods and Materials

### 2.1 Patient’s collection

Cohort I and cohort II included 1661 and 1678 cancer patients with ICI treatment acquired from previous studies, respectively [12, 15]. The clinical information of a without-ICI treatment cohort containing 25000 patients was acquired from cBioPortal databases (https://www.cbioportal.org/study/summary?id=msk_met_2021). The TMB values of all patients included in three cohorts were measured in the same way. Those cancer types shared by three cohorts were selected for subsequent analysis including bladder cancer, breast cancer, colorectal cancer, esophagogastric cancer, head and neck cancer, melanoma, non-small cell lung cancer, and renal cell carcinoma. Finally, the with-ICI treatment cohort contains 2767 patients, and the without-ICI treatment cohort contains 14862 patients.

### 2.2 Statistical analysis

Survival analysis was carried out using Kaplan–Meier curve, and the log-rank test was used to determine the statistical significance of differences. Survival analysis and univariate cox proportional hazard regression were used with OS and PFS as the endpoint. A *P* value of less than 0.05 was considered statistically significant, and all the hypothesis tests were 2-sided. All statistical analyses were conducted using R V.4.0.2.

## 3. Results

### 3.1 The landscape of TMB across cancer types

In the present study, we included eight types of tumors containing 2767 tumor patients who received ICI therapy and 14862 cases with tumors who did not receive ICI therapy, respectively (Figure 1, Table 1). As shown in Figure 2A, the median TMB of tumor patients with ICI treatment was quietly different from that in patients without ICI therapy, indicating that the definition of optimal cutoffs of TMB in a given tumor type in a previous study might own selection bias[15]. Therefore, we combined patients without ICI therapy with patients with ICI treatment to decrease selection bias in the following study. We tried to explore the possible cutoffs (≥10mut/Mb, top 10%, top 20%, top 30% of TMB within each histology) of TMB to predict prognosis and ICI response. The landscape and different cutoffs of TMB across cancer types was shown in Figure 2B.

**Figure 1.**
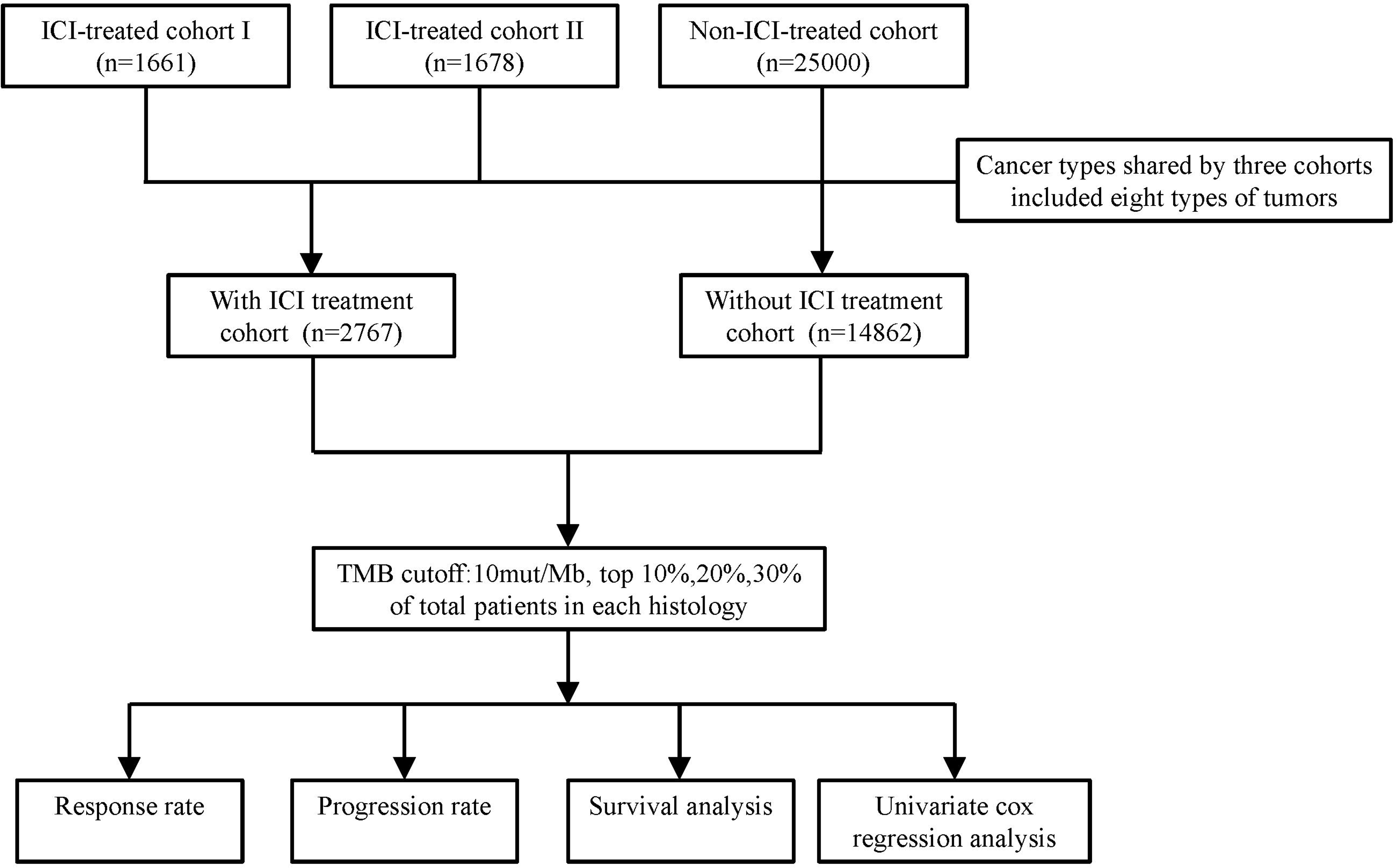
A diagram for the design of the present study.

**Figure 2.**
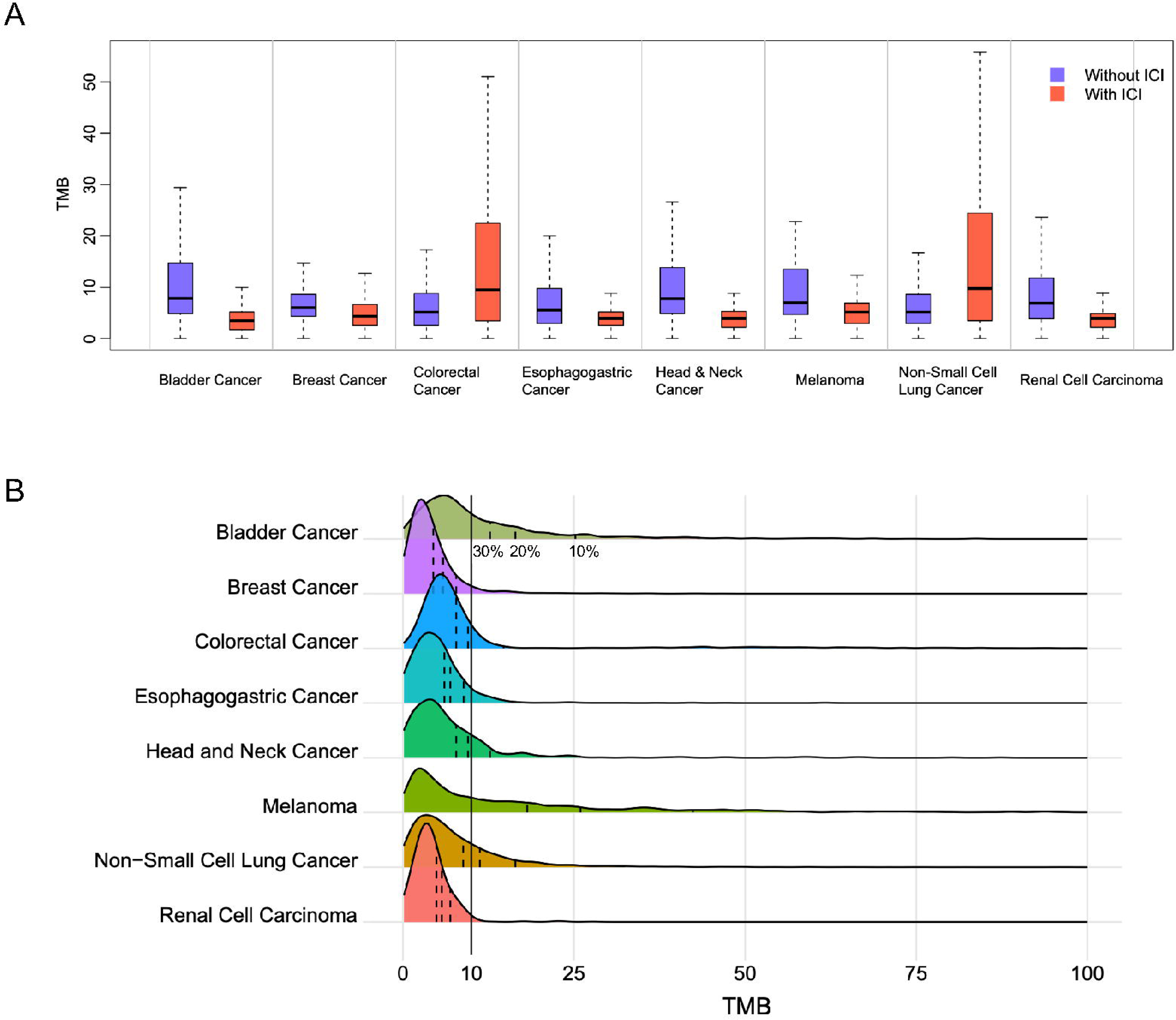
The landscape and different cutoffs of TMB across cancer types. A. The range of TMB distribution in ICI-treated and non-ICI-treated cohorts. B. Different cutoffs of TMB chosen in cancer types. Dotted lines mean top 10%, 20%, 30% of TMB in corresponding types of tumors from the right to left in turn. Solid line means 10 mut/Mb of TMB in cancers.

### 3.2 Association between TMB and response rate, progression rate, and survival rates

It’s generally accepted that high TMB generally predicts a favorable ICI response rate, however, we still don’t know whether there is a relationship between TMB and therapeutic response that the higher value of the TMB, the better the therapeutic response. As shown in Figure 3A, the response rates increased along with increased TMB cutoffs within cancer types. Cumulative response rates in patients with TMB lower than 5mut/Mb was only 22%, while reached 80% in those higher than 50mut/Mb. The TMB cutoff of 50% of cumulative response is about 15mut/Mb. In contrast, cumulative progression rates decreased following increased TMB cutoffs in total cancer types. The TMB cutoffs of 50% of the cumulative progression rate was about 25mut/Mb (Figure 3B). Survival rates were also positively correlated with TMB cutoffs, and the TMB cutoffs of 50% of the cumulative survival rate was about 10mut/Mb (Figure 3C). These results indicated that TMB cutoffs appropriate for predicting response rates, progression rates as well as survival rates were likely varied in ICI therapy patients.

**Figure 3.**
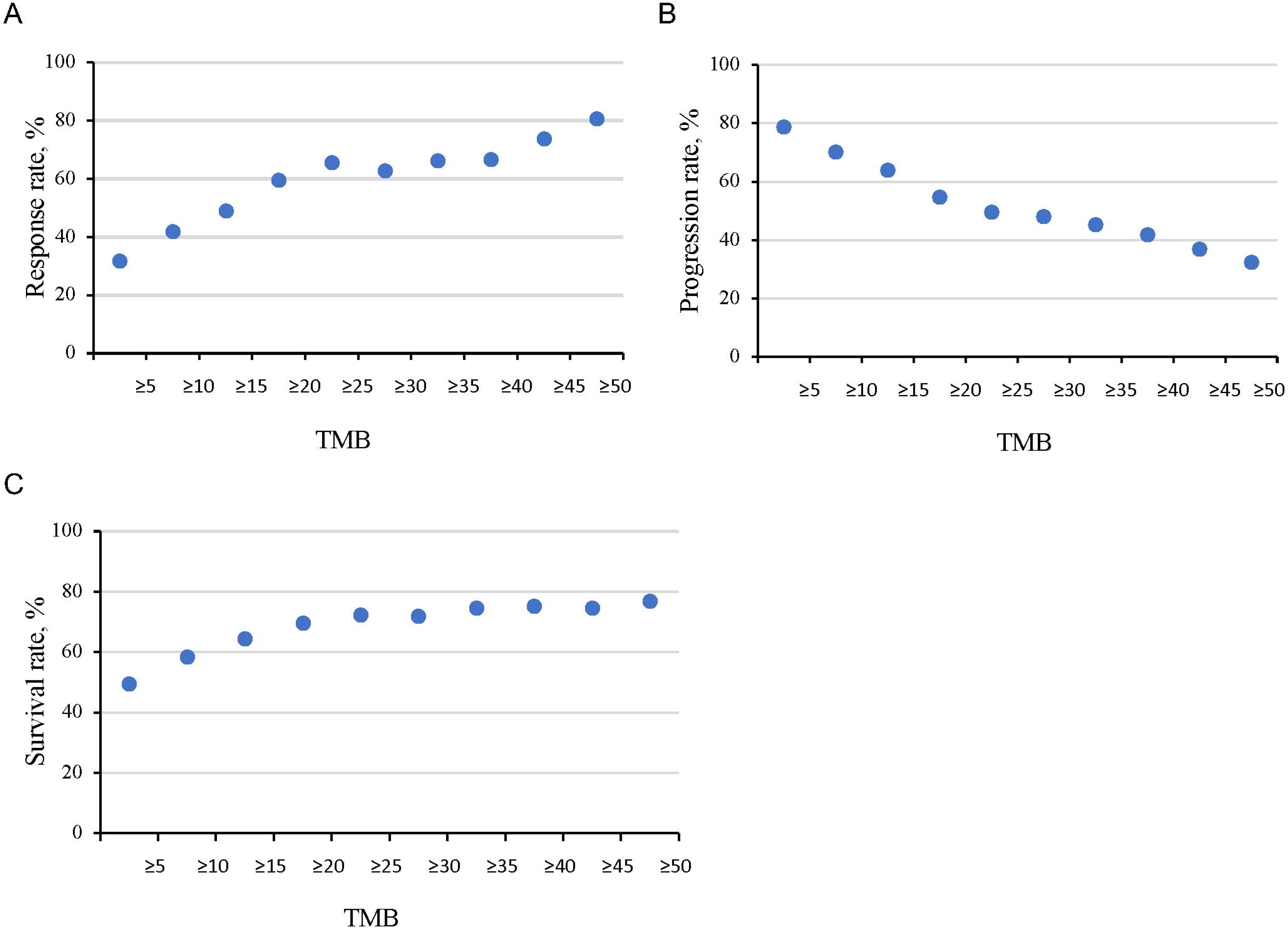
Association between TMB and response rate (A), progression rate (B), and survival rates(C).

### 3.3 Outcomes based on a universal TMB cutoff of 10 mut/Mb

Previous studies have shown that TMB≥ 10mut/Mb was associated with a favorable objective response and longer survival times with pembrolizumab monotherapy across several types of tumors including anal, biliary, cervical, endometrial, mesothelioma, neuroendocrine, salivary, small-cell lung, thyroid and vulvar cancer[16]. We first assessed the prognostic value of TMB (≥ 10 mut/Mb) across eight types of tumors. Among eight types of tumors, the TMB of all patients with renal cell carcinoma exhibited was lower than 10 mut/Mb, indicating that TMB≥ 10 mut/Mb is inappropriate for this type of tumor. As shown in Figure 4, patients with TMB≥ 10 mut/Mb present higher response rates and lower progression rates than that in TMB low groups in patients with bladder cancer, breast cancer, melanoma, and non-small cell lung cancer. In all patients receiving ICI treatment, patients with TMB ≥ 10 mut/Mb exhibited better OS and PFS than that with lower TMB (Figure 5A, C). In contrast, TMB presents no values for predicting OS in patients who did not receive ICI (Figure 5E). We further explore whether TMB≥ 10 mut/Mb is a valuable indicator for a given type of tumor. We found that patients with TMB≥ 10 mut/Mb were a favorable predictor for OS in five (bladder cancer, colorectal cancer, esophagogastric cancer, melanoma, and non-small cell lung cancer) out of seven types of tumors (Figure 5B). Besides, TMB ≥ 10 mut/Mb was also positively associated with PFS in patients with bladder cancer, melanoma, and non-small cell lung cancer (Figure 5D). However, we observed that patients with TMB≥ 10 mut/Mb also predicted favorable prognosis in bladder cancer, colorectal cancer, and melanoma without ICI treatment (Figure 5F). TMB≥ 10 mut/Mb was positively correlated with OS in both non-ICI and ICI therapy patients, which could not reflect whether ICI therapy was efficient for these three types of tumors. Taken together, these results indicated that TMB≥ 10 mut/Mb is not an ideal predictor for immunotherapy for all cancer types.

**Figure 4.**
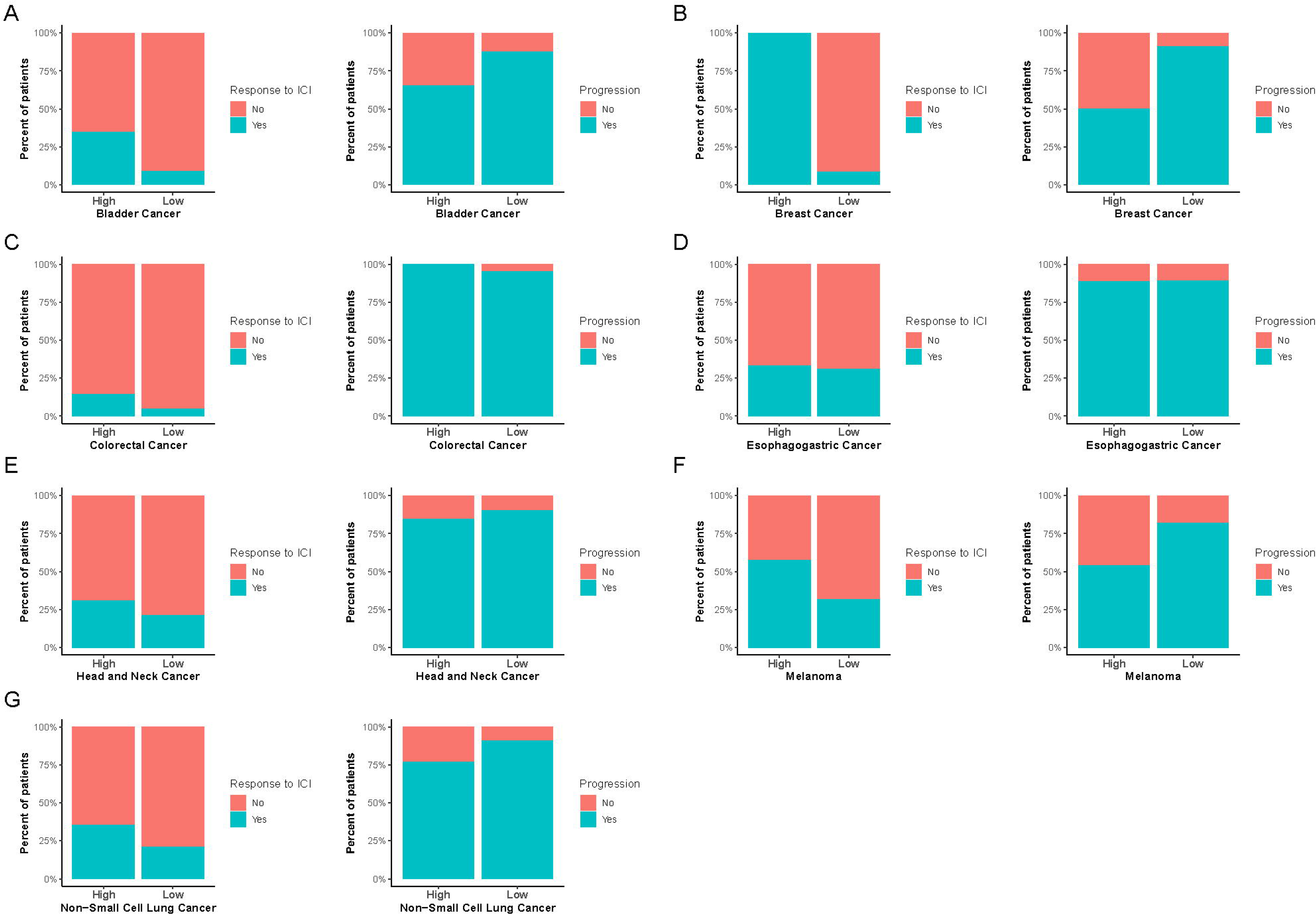
Response rates and progression rates in high- and low-TMB group in the ICI-treated cohort based on the cutoff 10 mut/Mb (A-C).

**Figure 5.**
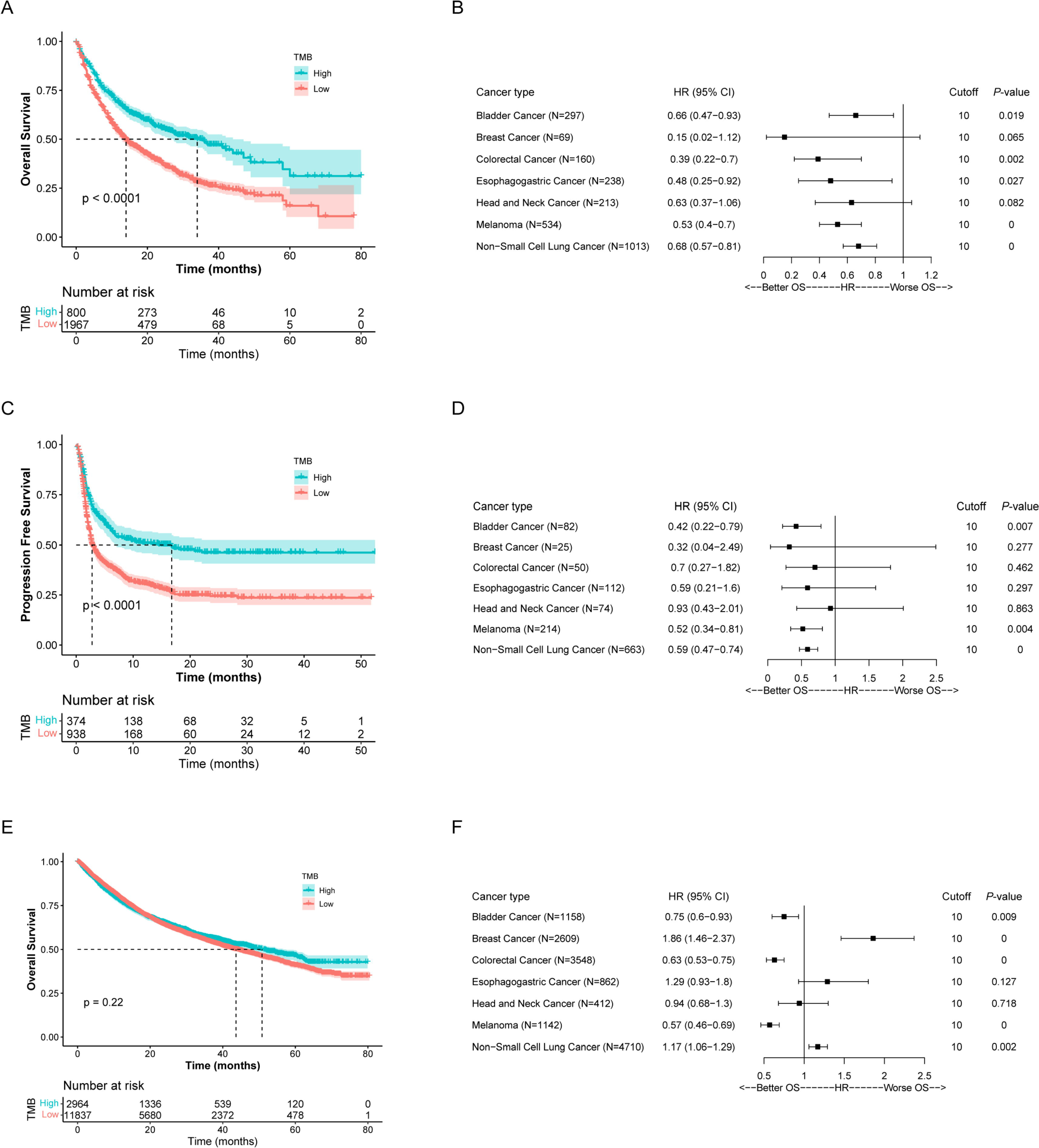
Correlation between TMB values and prognosis in patients in ICI-treated and non-ICI-treated cohorts based on the cutoff 10 mut/Mb. A-B Overall survival and cox analysis of TMB in cancers receiving ICI treatment. C-D. Progression survival and cox analysis of TMB in cancers receiving ICI treatment. E-F. Overall survival and cox analysis of TMB in cancers without ICI treatment.

### 3.4 Outcomes based on the top 10% of TMB in each cancer

The top 10% of TMB in bladder cancer, breast cancer, colorectal cancer, esophagogastric cancer, head, and neck cancer, melanoma, non-small cell lung cancer, and renal cell carcinoma was 25.94, 7.78, 27.67, 8.87, 12.97, 46.97, 16.42, and 6.91 mut/Mb, respectively. As shown in Figure 6A, patients with higher TMB exhibited a better response rate in bladder cancer, breast cancer, head and neck cancer, melanoma, non-small cell lung cancer, and renal cell carcinoma, while only presented a lower progression rate in melanoma, non-small cell lung cancer, and renal cell carcinoma concerning that with lower TMB. Patients with higher TMB only predicted longer PFS in melanoma, non-small cell lung cancer, and renal cell carcinoma (Figure S1). Higher TMB predicted longer OS in bladder cancer, colorectal cancer, melanoma, and non-small cell lung cancer who received ICI therapy. In patients without ICI therapy, higher TMB predicated poorer OS in breast and esophagogastric cancer, while predicted better OS in bladder cancer, colorectal, and melanoma (Figure 7).

**Figure 6.**
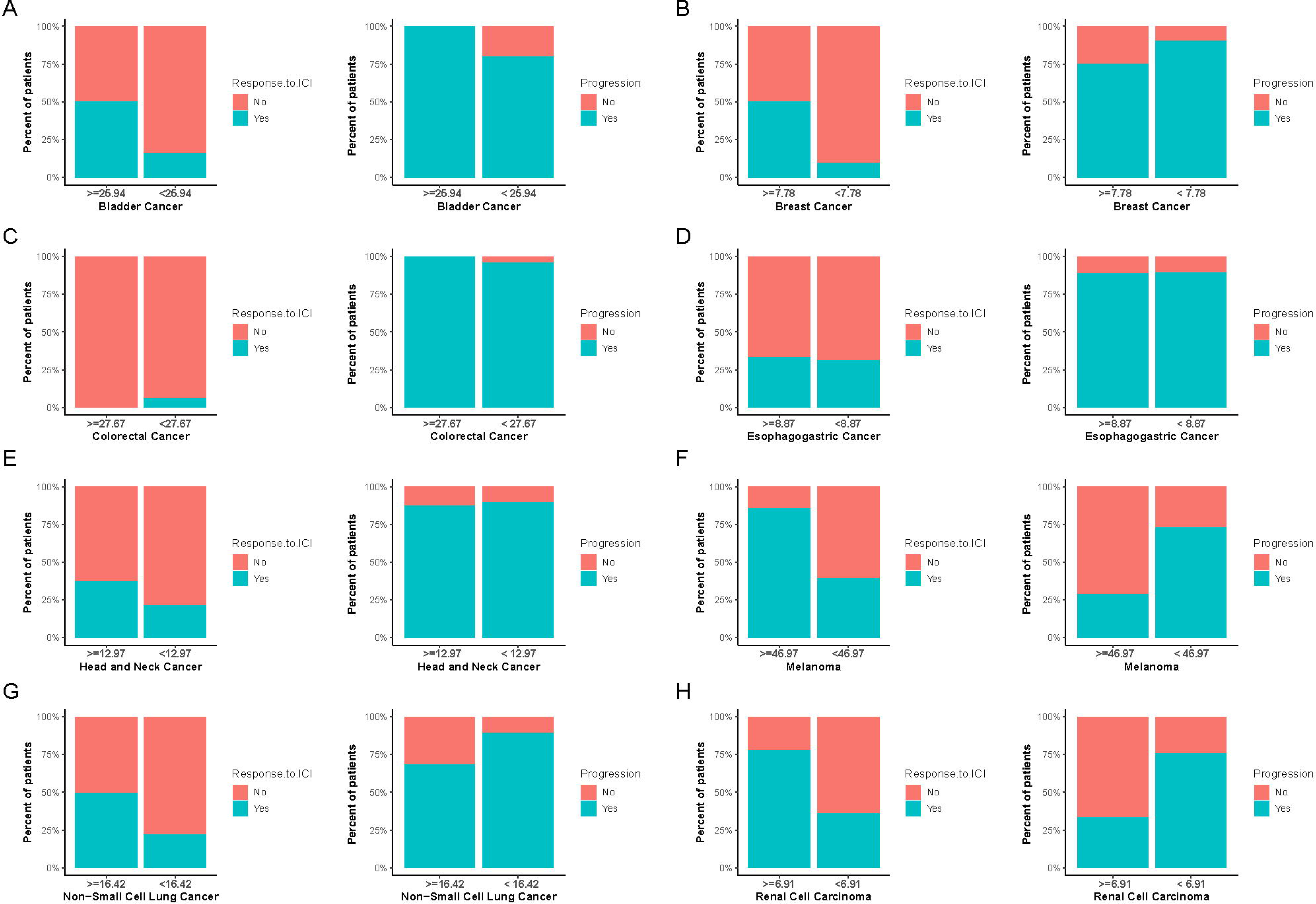
Response rates and progression rates in high- and low-TMB group in the ICI-treated cohort based on the cutoffs of top 10% (A-H).

**Figure 7.**
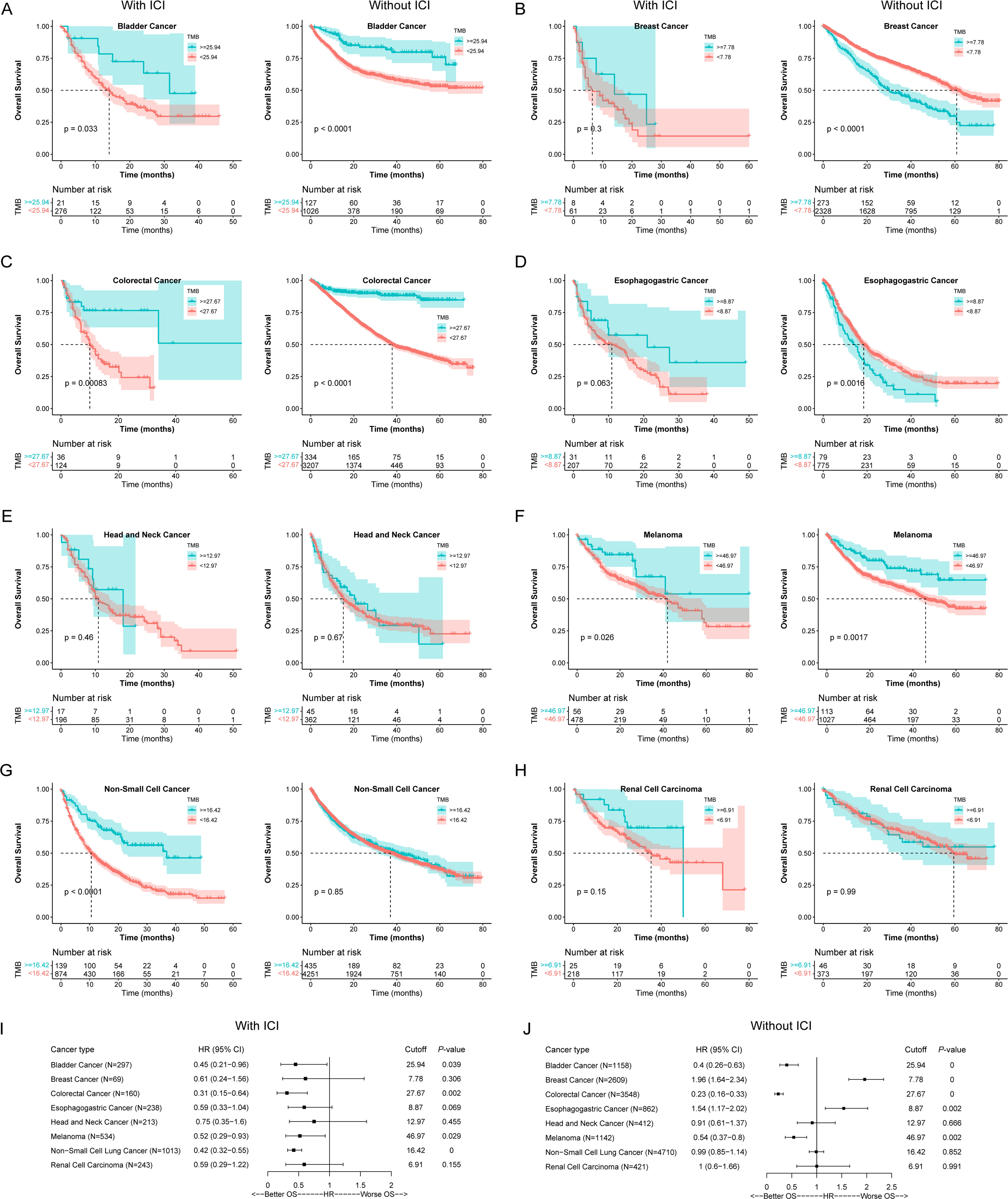
Kaplan-Meier survival (A-H) and cox analysis (I-J) of TMB in cancers receiving ICI treatment based on the cutoffs of top 10%.

### 3.5 Outcomes based on the top 20% of TMB in each cancer

The top 20% of TMB also varied from different types of tumors. As shown in Figure 8, response rates were reversely associated with progression rates in five (bladder cancer, breast cancer, head and neck cancer, melanoma, non-small cell lung cancer, and renal cell carcinoma) out of eight types of tumors. Patients with the top 20% of TMB exhibited longer PFS in bladder cancer, melanoma, and non-small cell lung cancer compared to that in patients with the bottom 80% of the TMB group (Figure S2). Besides, patients with the top 20% of TMB presented a better prognosis independent of ICI therapy in bladder cancer, colorectal cancer, and melanoma. Interestingly, in breast cancer, esophagogastric cancer, and non-small cell lung cancer, higher TMB predicted a poorer prognosis in non-ICI therapy patients (Figure 9).

**Figure 8.**
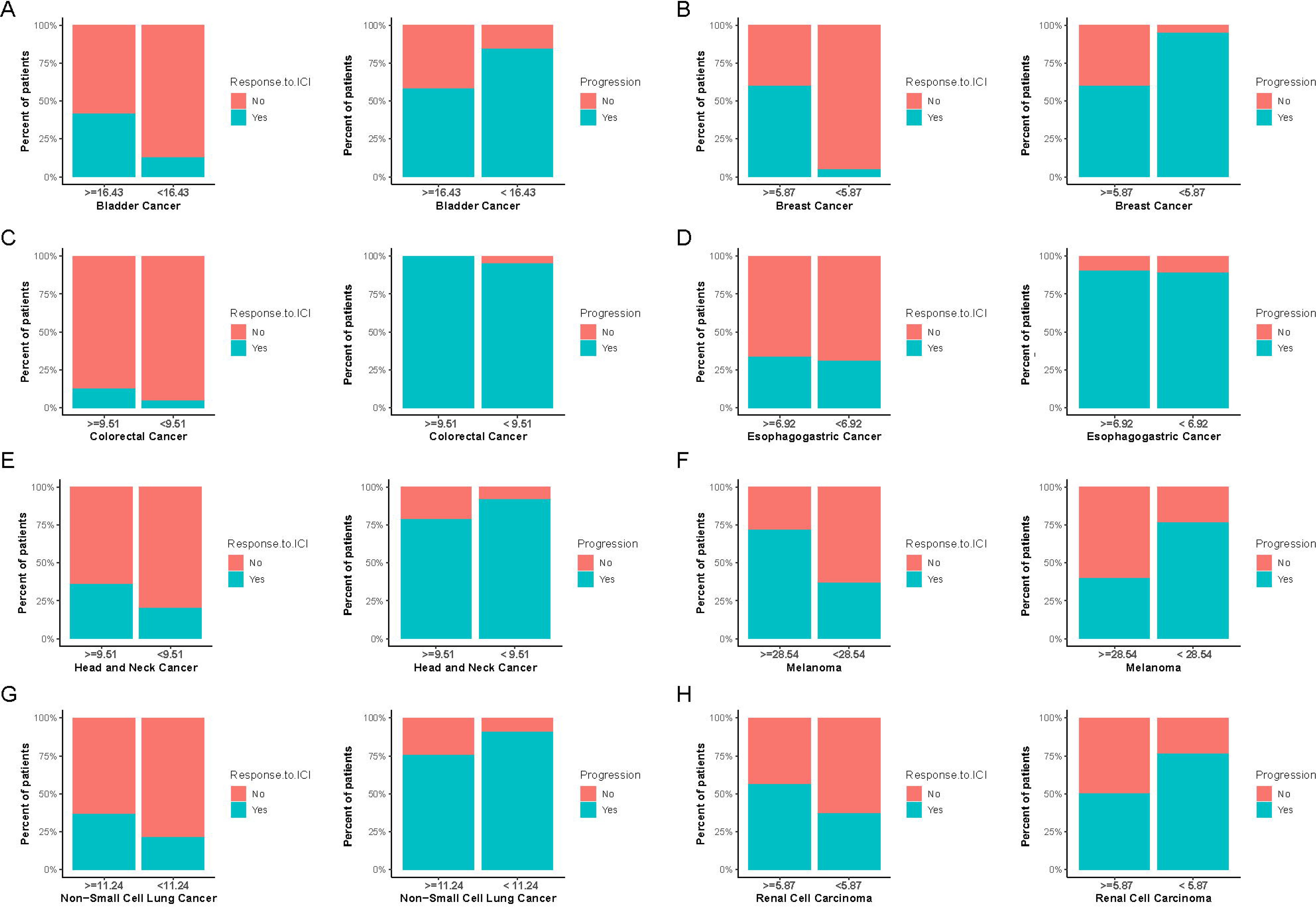
Response rates and progression rates in high- and low-TMB group in the ICI-treated cohort based on the cutoffs of top 20% (A-H).

**Figure 9.**
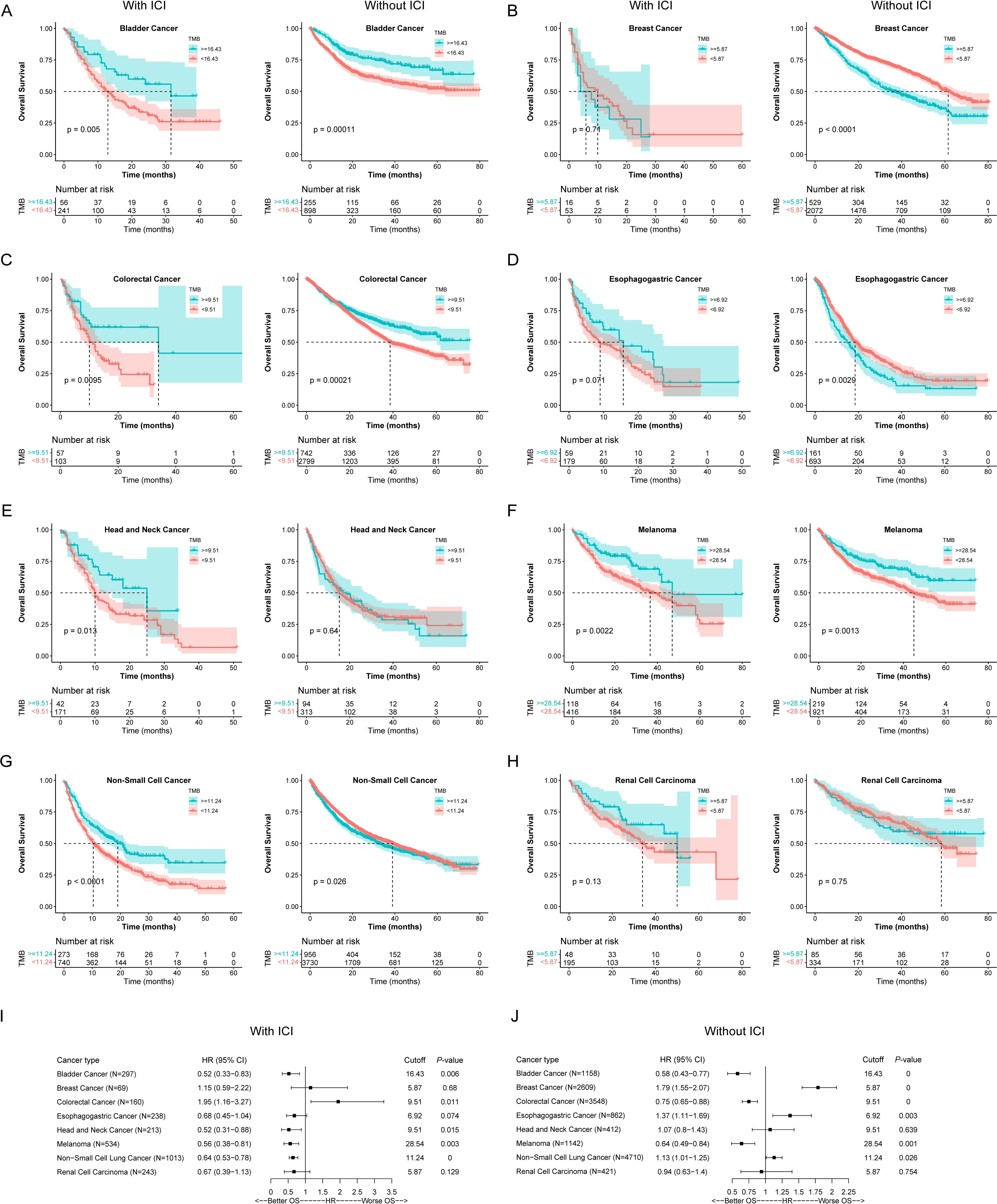
Kaplan-Meier survival (A-H) and cox analysis (I-J) of TMB in cancers receiving ICI treatment based on the cutoffs of top 20%.

### 3.6 Outcomes based on the top 30% of TMB in each cancer

We further explored the prognostic roles of the top 30% of TMB in cancers. Similar to that in the top 20% of TMB, response rates were reversely associated with progression rates in five (bladder cancer, breast cancer, head and neck cancer, melanoma, non-small cell lung cancer, and renal cell carcinoma) out of eight types of tumors(Figure S3). This cutoff is only associated with OS in four out of eight types of tumors with ICI therapy (Figure S4). This means that this result of survival analysis revealed that the prognostic roles of TMB in bladder and melanoma are independent of ICI therapy. Despite that, the top 30% of TMB predicted better PFS across multiple cancer types (Figure S5). The remaining three types of tumors did not reach significant statistics partially due to smaller sample size.

## 4. Discussion

In the Keynote-158 trial, the researchers reported that TMB≥10 mut/Mb acted as a meaningful predictive biomarker for response to pembrolizumab monotherapy in ten types of tumors[16]. Based on the above results, the FDA has approved pembrolizumab for the immunotherapy of adult and pediatric solid tumors of any histologic type with TMB≥10 mut/Mb. However, we found that the optimal cutoffs of TMB in five out of eight types of tumors were lower than 10mut/Mb in our present study (Table 2). Notice that, all patients included in the Keynote-158 trial with high microsatellite instability (MSI-H), which tend to increase the values of TMB [16–18]. In contrast, patients enrolled in our present study were all without MSI-H. Moreover, the TMB values in primary tumors were lower than that in metastatic tumors[19]. Nearly 60% of total patients included in the present study were primary tumors, while all patients included in the Keynote-158 trial were unresectable or metastatic tumors[16]. The above findings might partially account for the lower optimal cutoffs of TMB in most types of tumors in the present study.

Bladder cancer ranks as the most common malignant tumor of the urinary system, accounting for nearly 170,000 deaths worldwide [20]. The wide mutational profiles containing antigenic enable patients with bladder cancer benefits from immunotherapies [21, 22]. In this study, we found that patients with top 20% or 30% of TMB or TMB≥10 mut/Mb exhibited higher response rates and lower progression rates compared with the cutoffs of top 10% of TMB. Besides, patients with TMB≥10 mut/Mb also presented longer PFS. Therefore, TMB≥10 mut/Mb might serve as ideal cutoffs for predicting response rate, progression rates, and progression-free survival. Interestingly, we noticed that higher TMB could predict better OS in both ICI and non-ICI therapy patients, indicating that TMB was inappropriate for predicting OS in ICI therapy patients. However, TMB≥10 mut/Mb could serve as a favorable indicator for patients without ICI therapy.

Breast cancer is one of the most common cancers worldwide and accounts for about 30% of female cancers[23]. Up to now, many ICI drugs were used in the treatment of breast cancer[24]. It’s reported that high TMB(≥10mut/Mb) was correlated with improved PFS but not OS in patients with metastatic triple-negative breast cancer who received PD-1 checkpoint treatment[25]. Nevertheless, we found that TMB≥10mut/Mb did not correlate with PFS in our present study. Instead, we found that only the top 30% of TMB(≥4.44mut/Mb) was associated with higher response rates, lower progression rates, and increased PFS. However, compared with the top 30% of TMB, it seemed that the top 20% of TMB (≥5.87mut/Mb) were more efficient for predicting response rates and progression rates. Similar to the previous study, we also observed no statistical correlation between TMB and OS in patients receiving ICI treatment[25]. Interestingly, high TMB(≥4.44mut/Mb) could predict poor prognosis in patients without ICI therapy.

Colorectal cancer is a leading cause of cancer-related death worldwide and the third most common cause of cancer mortality worldwide with more than 1.85 million cases and 850[000 deaths annually[26, 27]. In the immunotherapy of colorectal cancer, nivolumab was the first immune checkpoint inhibitor approved by FDA[28]. Data from an open-label, clinical trials suggested that patients, who received ICI treatment, with high TMB (between 37 and 41 must/Mb) exhibited longer PFS[29]. Results from a meta-analysis showed that the optimal cutoffs of TMB for predicting prognosis varied from 10 to 96 mut/Mb[30]. In contrast, we found that cutoffs of TMB≥7.82 mut/Mb(top 30%) were associated with both increased PFS and longer OS.

Esophagogastric cancer represents a significant global health problem, with most patients presenting with advanced-stage disease and consequently with a poor prognosis[31]. Despite the uses of immune checkpoint inhibitors have now been advanced to include the first-line treatment of esophagogastric cancers, only one study has explored the correlation between TMB and survival times [32, 33]. In that study, there was no significant relationship between TMB as a continuous variable and PFS or OS in multivariable analysis. However, when TMB was categorized by quartiles, high TMB(≥7.82 mut/Mb) was associated with improved PFS[33]. Although we did no observe different response and progression rates between high and low-TMB groups regardless of the four cutoffs(≥10 mut/Mb, top 10%, or 20%, or 30% of TMB), there indeed was a significant correlation between high TMB and better prognosis. We found that patients receiving ICI treatment with TMB≥10 mut/Mb were associated with longer OS, while TMB≥6.92 mut/Mb (top 20%) was correlated with poor OS in patients without ICI therapy. Nevertheless, we did not find an appropriate cutoff of TMB for predicting PFS in patients with ICI therapy.

Head and neck cancer contains multiple subtypes of tumors and ranks as the seventh most common type of cancer [34]. Immunotherapy has changed the therapeutic options in medical treatments of head and neck cancer and TMB has been an explored biomarker. In the immunotherapy of head and neck cancer, it’s reported that TMB was higher in responders compared with non-responders[35]. However, results from another ICI cohort containing 261 patients presented that TMB≥10 mut/Mb failed to predict both OS and disease-free survival[36]. Our present study demonstrated that the top 20% of TMB (≥9.51 mut/Mb) could predict OS in patients with ICI treatment, at which cutoff, TMB exhibited no predictive role for that without ICI therapy. However, no significant correlation between TMB and PFS was observed.

Genetic inter- and intra-tumor heterogeneity of tumors made malignant melanoma notorious[37]. Despite that, immune checkpoint inhibitors have successfully regressed tumor cell growth and nearly 50% of patients reached long-term durable cancer control concerning no more than 10% historically in advanced melanoma [38]. TMB≥16 mut/Mb is related to OS, and PFS in patients with atezolizumab, one kind of ICIs, treatment[39]. Our present study found that the cutoff of TMB≥10 mut/Mb was enough for predicting response rates, progression rates, PFS, and OS in patients with ICI therapy. Interestingly, anti-PD-1 immunotherapy was reported to lower TMB values, which, in turn, influence responses to adoptive cell transfer in anti-PD-1-experienced patients[40].

Non-small cell lung cancer, a subtype of lung cancer, accounts for 85% of all lung cancer[41]. Initial studies revealed that ICI treatment could effectively activate T cells to antitumor and patients could benefit from ICI therapy in phase I and II studies[42]. Among multiple biomarkers, TMB was an essential predictor for ICI therapy [43, 44]. The impact of DNA damage response and repair gene mutations on the efficacy of ICI therapy was partially due to the different distribution of TMB between mutation and non-mutation groups[45]. In this study, we found that TMB≥8.808 mut/Mb was positively correlated with PFS and OS in ICI therapy patients, while negatively associated with prognosis in patients without ICI treatment. These findings demonstrated that TMB was a valuable indicator for the prognosis of patients with non-small cell lung cancer.

The management of patients with metastatic renal cell carcinoma has changed dramatically and ICI therapy has been approved in 2017 [46]. Although TMB was positively correlated with the production of neoantigens, there was no significant correlation between TMB and prognosis in patients with ICI treatment in renal cell carcinoma [47, 48]. Similar to the previous study, we also found that TMB was not associated with PFS and OS in patients receiving ICI treatment. In combination with these results, we might conclude that TMB was not an ideal predictor for patients with ICI therapy in renal cell carcinoma.

## 5. Conclusion

Our present study tried to explore possible universally optimal cutoffs of TMB for predicting immunotherapy response and prognosis for eight types of tumors. We found that optimal cutoffs of TMB were in a tissue-dependent manner. Besides, optimal cutoffs for PFS and OS might also differ in a given type of tumor. Hence, we concluded that high tumor mutational load predicts survival and responses to ICI immunotherapy in a tissue-dependent manner. However, the optimal cutoffs of TMB for a given type of tumor in our present study are not all consistent with that in previous studies, the different methods of measurement for TMB may partially account for this[49]. In addition, cases in some types of tumors with ICI treatment included in the current study were limited. Further larger ICI cohorts, the TMB values of which are measured with the same method, are needed to provide more precise optimal cutoffs of TMB.

## Abbreviations

TMB: Tumor mutational burden
ICIs: Immune checkpoint inhibitors
PD-1: Programmed death 1
PD-L1: Ligand of the PD-1 axis
CTLA4: Cytotoxic T lymphocyte antigen 4
FDA: United States Food and Drug Administration
PFS: Progression-free survival
OS: Overall survival
MSI-H: high microsatellite instability

## Acknowledgements

Not applicable.

## Author contributions

Zhi-Qiang Li and Ze-Fen Wang were involved in the conception and the design of the study. Feng Tang, Tian Lan and Zhen-Yuan Liu performed bioinformatics analysis and participated in the acquisition and analyzation of the data. Feng Tang and Tian Lan wrote the manuscript. Zhi-Qiang Li and Ze-Fen Wang reviewed the manuscript. All authors reviewed the manuscript and approved the final version of the manuscript.

## Data availability

The data used to support the findings of this study are available from the corresponding author upon request.

## Declarations

### Ethics approval and consent to participate

Not applicable.

