## Supplementary material and Table1 Table2 for "High tumor mutational burden predicts survival and responses to ICI immunotherapy in a cancer-dependent manner"

**Figure legends**

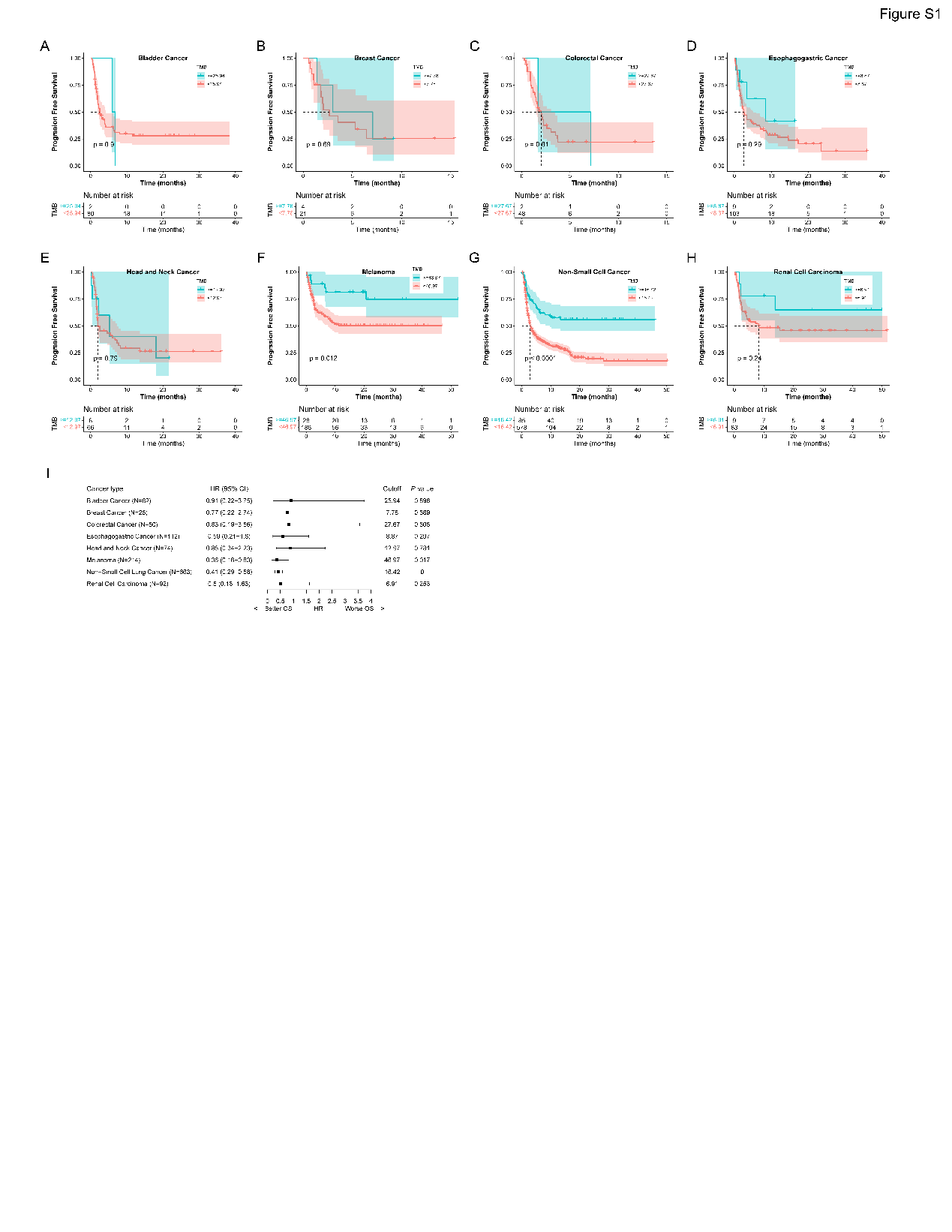

**Figure S1. Progression free survival (A-H) and cox analysis (I) of TMB in cancers receiving ICI treatment based on the cutoffs of top 10%.**

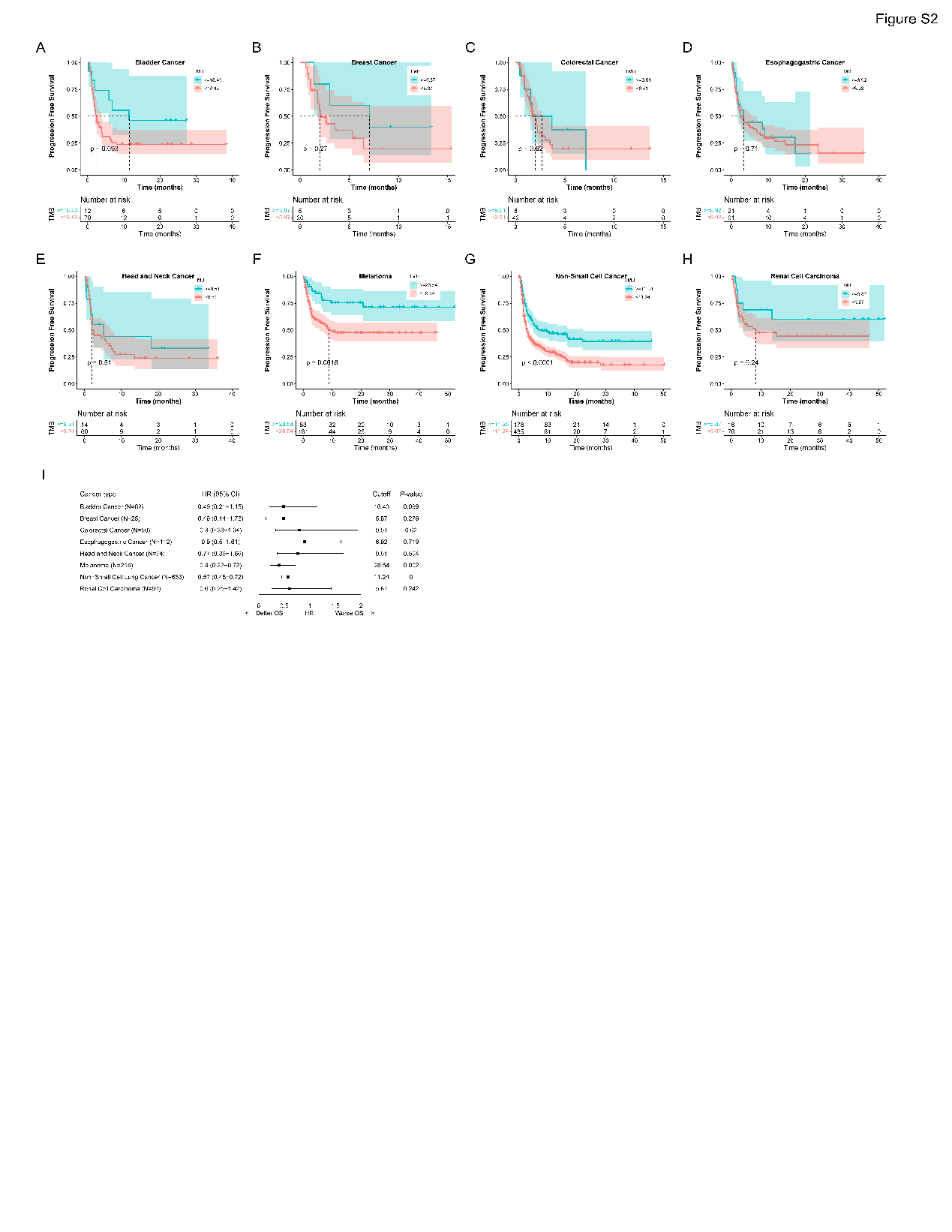

**Figure S2. Progression free survival (A-H) and cox analysis (I) of TMB in cancers receiving ICI treatment based on the cutoffs of top 20%.**

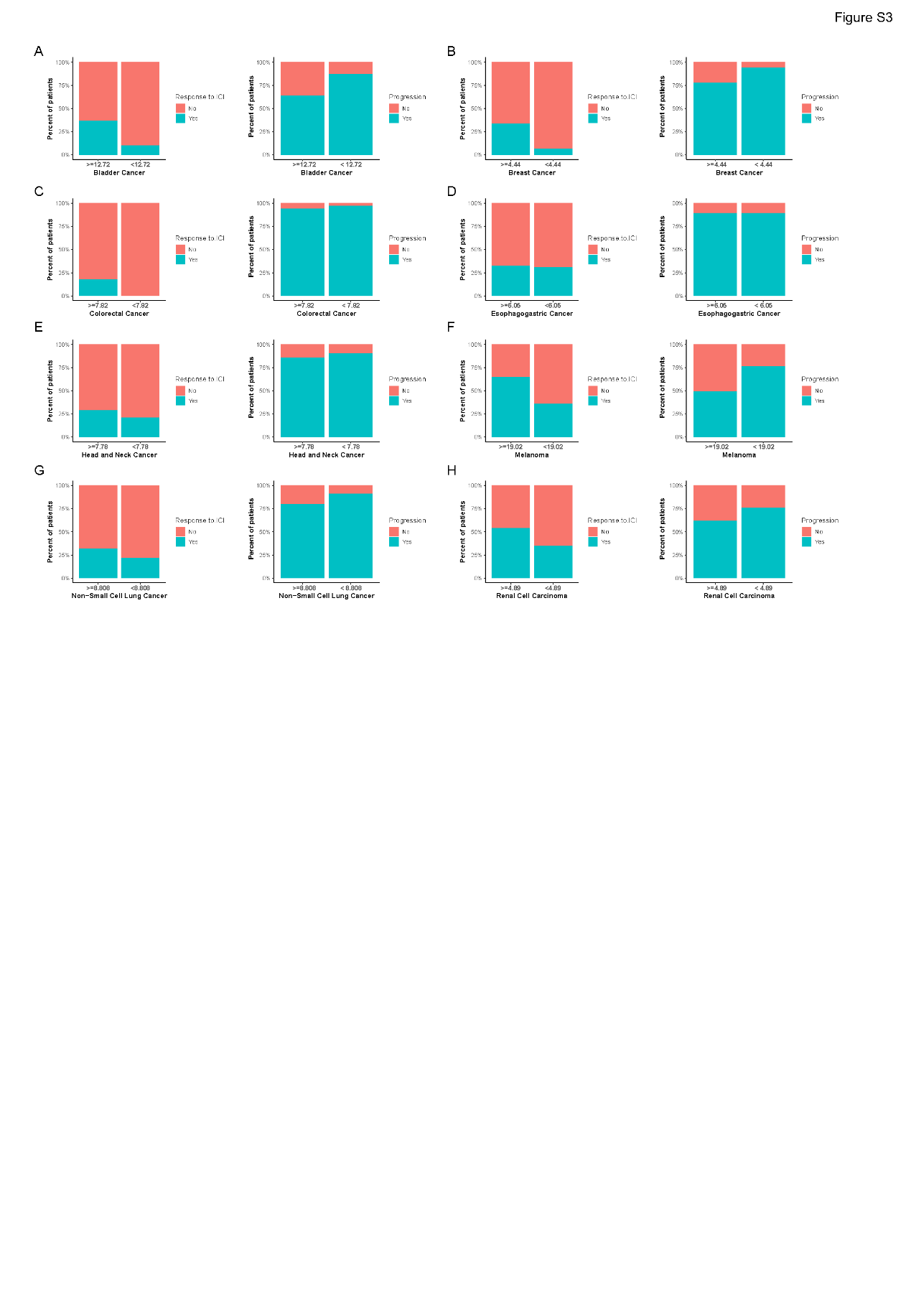

**Figure S3. Response rates and progression rates in high- and low-TMB group in the ICI-treated cohort based on the cutoffs of top 30% (A-H).**

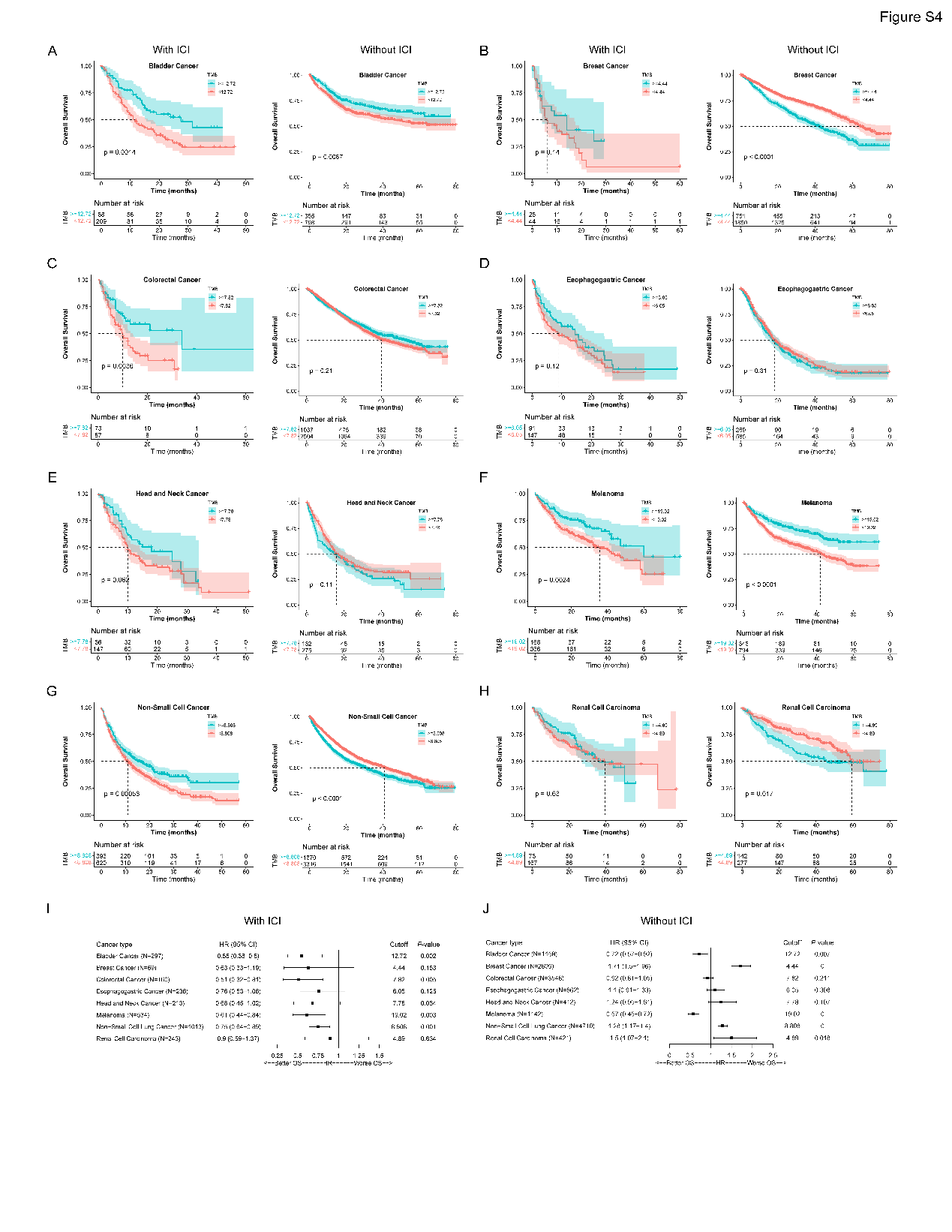

**Figure S4. Kaplan-Meier survival (A-H) and cox analysis (I-J) of TMB in cancers receiving ICI treatment based on the cutoffs of top 30%.**

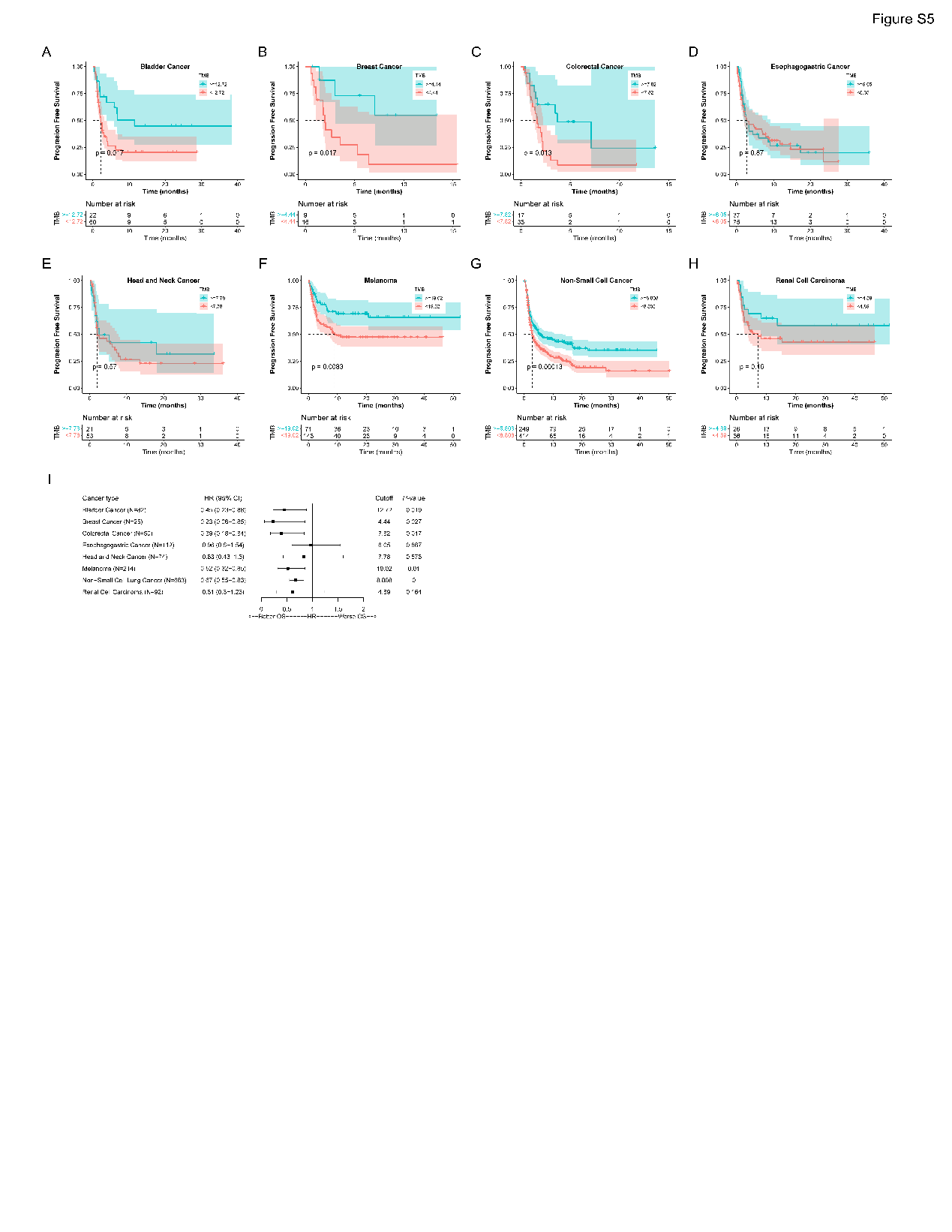

**Figure S5. Progression free survival (A-J) and cox analysis (I) of TMB in cancers receiving ICI treatment based on the cutoffs of top 30%.**
